# Epidemiological analysis of hospitalized cases of COVID-19 in indigenous people in an Amazonian region: cross-sectional study with data from the surveillance of acute and severe respiratory syn-dromes in Brazil

**DOI:** 10.1101/2021.12.15.21267872

**Authors:** Ana Lucia da Silva Ferreira, Daniele Melo Sardinha, Claudia Ozela El-Husny, Carmem Aliandra Freire de Sá, Emilyn Costa Conceição, Karla Valéria Batista Lima

## Abstract

Indigenous people are considered more vulnerable to new infectious agents. In view of the novel coronavirus causing COVID-19, health authorities are concerned about the possible impact of the pandemic on reaching vulnerable populations, such as the indigenous people of the Brazilian Amazon. Thus, we aimed to carry out an epidemiological analysis of serious cases and deaths from COVID-19 in indigenous population in the state of Pará, Brazil. The data was obtained from the public Ministry of Health platform. Data analysis was performed using the Statistical Package for the Social Sciences 20, Chi-square of adherence, the independence test and G test. For spatial distribution was used ArcGIS. We observed 123 COVID-19 cases: 46 deaths (37.40%), male gender (76-61.79%), age above 60 years (61-49.6%), the most frequent risk factor was chronic cardiovascular disease (18-14.63%). The predictors of death were: invasive ventilation has (10.73) more chances for the outcome death, those not vaccinated against influenza have (3.41) and age (1.4). COVID-19 occurrence was higher in municipalities that have villages with health care or commerce, or with migrants from the Warao ethnic group. Notifications should take into consideration the specific issues of indigenous people so that effective control measures can be defined.

## 1. Introduction

In view of the novel coronavirus causing COVID-19, health authorities are concerned about the possible impact of the pandemic on reaching vulnerable populations, such as the indigenous peoples of the Brazilian Amazon since studies have shown high rates of respiratory disease outbreaks in indigenous peoples. This concern increases since during the pandemic caused by H1N1 in 2009, the indigenous mortality rate was four times higher than the mortality of the general Brazilian population (Ribeiro & Rossi, 2020).

Indigenous peoples are considered more vulnerable to new infectious agents due to their genetic inability to respond immunologically to their initial contact with new infections. Linked to this, there are social, environmental, cultural and health factors such as high prevalence of tuberculosis and malaria, difficult access to clean water, malnutrition, and difficult implementation of control measures, such as social isolation (Amigo, 2020; Simionatto, Barbosa, & Marchioro, 2020; Vallinoto, da Silva Torres, Vallinoto, & Cayres Vallinoto, 2020).

The transmission of COVID-19 in the recent months in Brazil, resulted in an accelerated increase in the infection rate amongst the indigenous population in both rural and urban areas. It currently affects about 40,000 cases and 800 deaths (Pontes, Cardoso, Bastos, & Santos, 2021).. In the state of Pará, the first confirmed death from the disease occurred in an 87-year-old indigenous woman of the Borari ethnic group who lived in the village of Alter do Chão in Santarém (Cavalcante, 2020).

In this scenario, understanding the spatial distribution of cases and deaths by COVID-19 is essential to develop disease control strategies, allowing the identification of the main focuses, related comorbidities and possible implementation of disease control and treatment policies, such as awareness of social isolation and prevention measures and implementation of campaign hospitals for this population. Thus, we aimed to carry out an epidemiological analysis of serious cases and deaths from COVID-19 in indigenous population in the state of Pará.

## 2. Materials and Methods

### 2.1 Type of study

This is a cross-sectional, analytical, and ecological epidemiological study, based on data of severe cases and deaths from COVID-19 among indigenous population from the state of Pará. The data was obtained from the public web platform OpenDataSUS (https://opendatasus.saude.gov.br/) of the Ministry of Health and DataSUS (https://datasus.saude.gov.br/), which are related to the surveillance of the Acute and Severe Respiratory Syndromes of the Flu Epidemiological Surveillance Information System (*Síndromes Respiratórias Agudas e Graves do Sistema de Informação de Vigilância Epidemiológica da Gripe / SIVEP-Gripe*). This study followed the recommendations of the ‘The Strengthening the Reporting of Observational Studies in Epidemiology (STROBE) Statement: guidelines for reporting observational studies’ (von Elm et al., 2007).

### 2.2 Research location

Pará is a Brazilian state located in the North region, with the city of Belém as its capital. It is the second largest unit of the federation after Amazonas, with an area of 1.24 million km². The state belongs to the Amazon biome, and because of this the Equatorial climate predominates, besides the vegetation cover formed by forests and cerrados in a small part to the south. The Paraense economy, led by mineral and vegetal extraction, is the largest in the North region. Population: 8,690,745 inhabitants (2020 estimate), Demographic Density: 6.07 inhabitants/km² (IBGE, 2021).

### 2.3 Selection of participants

Severe Acute Respiratory Syndrome (SARS) is defined for individuals with Influenza Syndrome (IS) who have: dyspnea/respiratory distress OR persistent chest pressure OR O2 saturation less than 95% on room air OR bluish coloration of the lips or face. (IS: Individual with an acute respiratory condition characterized by at least two (2) of the following signs and symptoms: fever (even if referred), chills, sore throat, headache, cough, runny nose, smell or taste disturbances). For the purpose of notification in SIVEP-GRIPE, hospitalized cases of SARS or deaths from SARS regardless of hospitalization should be considered (M. da S. Brasil, 2020).

As the inclusion criteria, were selected confirmed cases for COVID-19, residents of Pará, indigenous race, notified from March 1, 2020, to February 22, 2021; and as the exclusion criteria, we selected the open cases with without case confirmation or unknown case evolution (figure 1).

**Figure 1.**
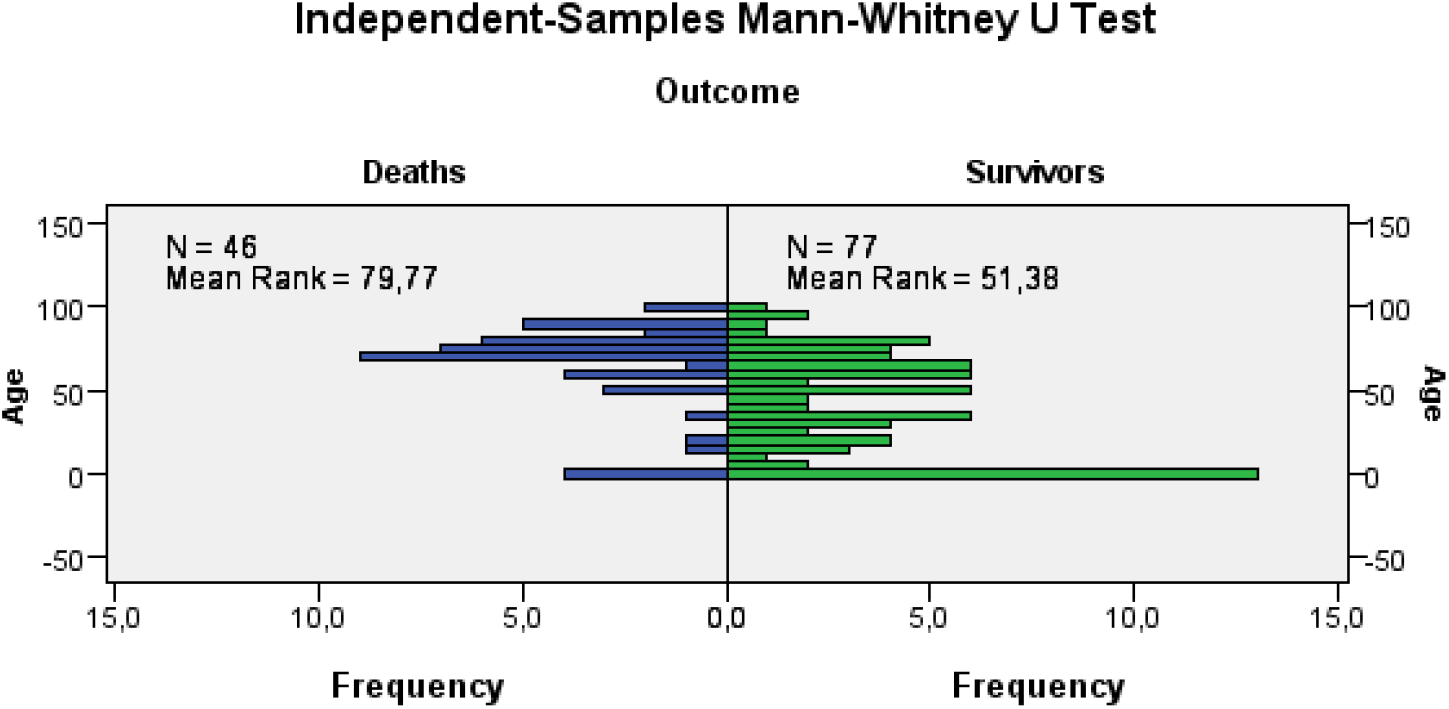
Age and outcome between deaths versus survivors in SARS by COVID-19 in indigenous people, 03/01/2020 to 02/22/2021, Pará, Brazil. Mann-Whitney U 954,500/Wilcoxon W 3.956,500/Test Statistic 954,500/Standart error 191,166/Standardized test statistic -4,276/Asymptotic Sig. (2-sided test) p-value <0.01. Source: OpenDataSUS, Ministry of Health, Brazil.

**Figure 1.**
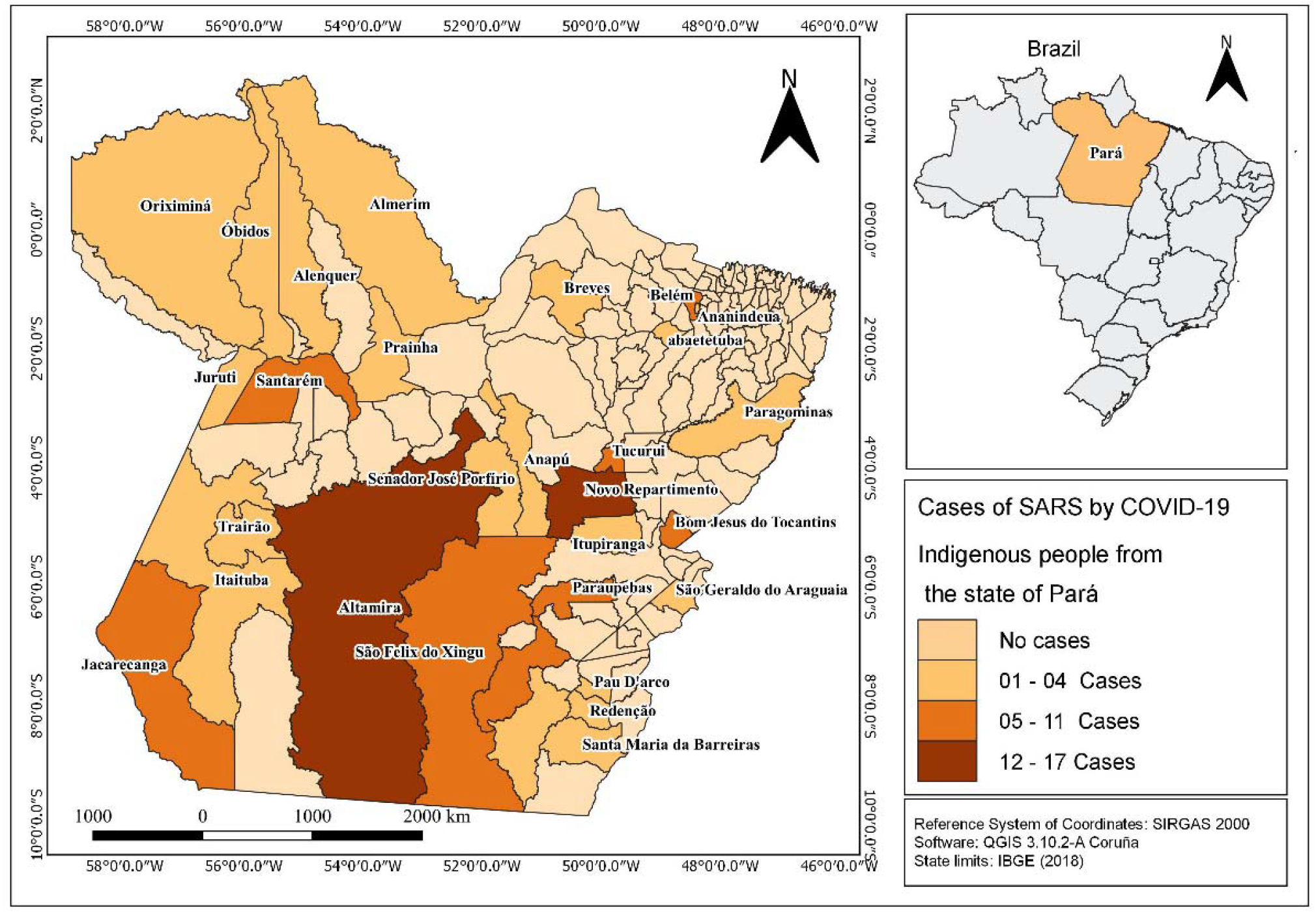
Cases of SRAG by COVID-19 in indigenous people, by municipality, 03/01/2020 to 02/22/2021 Pará, Brazil. Source: OpenDataSUS, Ministry of Health, Brazil.

### 2.4 Data Collection and Analysis

The database was available in Microsoft Excel 2019 format, and the variables were selected based on the SIVEP-Gripe notification form (M. da S. Brasil, 2020), consisting of 80 variables, referring to sociodemographic and clinical-epidemiological data. The variables extracted were sex (item 8), age (item 10), race/color (item 12), municipality of residence (item 19), signs and symptoms (item 35), have risk factors/comorbidities (item 36), took flu vaccine (item 37), was admitted to the intensive care unit (ICU) (item 47), used ventilatory support (item 50), final classification (item 72), termination criteria (item 73), and evolution (item 74).

Data analysis was performed using the Statistical Package for the Social Sciences 20 (SPSS – https://www.ibm.com/analytics/spss-statistics-software). From the Chi-square statistical tests of adherence and the independence test and G test (Contingency Table L x C) at values equal to or <0.05, to associate the significant variables between survivors and deaths. The results were presented in tables.

The Mann-Whitney test was applied to assess age differences between survivors and deaths. The multivariate logistic regression test was applied to assess the dependence of the death variable on the covariate’s invasive ventilation; not vaccinated against influenza; admitted to ICU, which were significant in the G-test, Sex and age were also included to see if age and sex could be a confounding factor. For all tests, an alpha level of significance of 0.05 was considered. The spatial distribution was performed in ArcGIS software (https://www.arcgis.com/) using the number of cases by municipality of residence and the quartile to separate the classes, without cases (0) – (1-4) – (5-11) and (12-17).

### 2.5 Ethical Aspects

The data of this study were made publicly available, not containing personal data of patients such as name, address, and telephone contact, thus, not presenting risks to the research participants, as well as the ethical opinion being dispensed with. This study is in accordance with Law No. 12,527, of 11/18/2011 (Access to Information Law) (M. da J. Brasil, 2011).

## 3. Results

### 3.1. Description of clinical and epidemiological data

The study population resulted in 123 cases of SARS per COVID-19 and 46 deaths (37.40%), mostly male (76-61.79%), aged over 60 years (61-49.60 %), the most frequent risk factor is chronic cardiovascular disease (18-14.63%). In addition to these, the laboratory confirmation criteria for analyzing the diagnosis of this population were evaluated (111-90.24%) (Table 1).

**Table 1.** Epidemiological profile of SARS by COVID-19 in indigenous peoples in the state. 03/01/2020 to 02/22/2021, Pará, Brazil.

| Epidemiological Characterization | Number | % | p-value <sup>(1)</sup> |
| --- | --- | --- | --- |
| **Age Range (year)** |  |  | <0.01 |
| < 1 | 13 | 10.57 |  |
| 01 to 19 | 13 | 10.57 |  |
| 20 to 39 | 17 | 13.82 |  |
| 40 to 59 | 19 | 15.45 |  |
| 60 to 79 | 42 | 34.15 |  |
| ≥ 80 | 19 | 15.45 |  |
| Total | 123 | 100 |  |
| **Sex** |  |  | <0.01 |
| Female | 47 | 38.21 |  |
| Male | 76 | 61.79 |  |
| Total | 123 | 100 |  |
| **Signs and symptoms** |  |  | <0.01 |
| Fever | 94 | 76.42 |  |
| Cough | 93 | 75.61 |  |
| Sore throat | 41 | 33.33 |  |
| Dyspnoeal | 90 | 73.17 |  |
| Respiratory discomfort | 79 | 64.23 |  |
| O2 saturation <95% | 70 | 56.91 |  |
| Diarrheal | 17 | 13.82 |  |
| Vomit | 12 | 9.76 |  |
| Total | 123 | 100 |  |
| **Risk factors** |  |  | <0.01 |
| Chronic Cardiovascular Disease | 18 | 14.63 |  |
| Asthma | 1 | 0.81 |  |
| Diabetes mellitus | 10 | 8.13 |  |
| Chronic Neurological Disease | 2 | 1.63 |  |
| Other Chronic Lung Disease | 1 | 0.81 |  |
| Immunodeficiency / Immunodepression | 2 | 1.63 |  |
| Chronic Kidney Disease | 4 | 3.25 |  |
| Obesity | 3 | 2.44 |  |
| Total | 123 | 100 |  |
| **Confirmation Criteria** |  |  | <0.01 |
| Laboratorial | 111 | 90.24 |  |
| Clinical-epidemiological | 1 | 0.81 |  |
| Clinical | 3 | 2.44 |  |
| Clinical image | 8 | 6.50 |  |
| Total | 123 | 100 |  |

<sup>1</sup> Adhesion test. Source: OpenDataSUS, Ministry of Health, Brazil.

### 3.2. Deaths versus survivors analysis

In the Mann-Whitney Test, death was associated with age (<0.0001). Therefore, it is understood that the older the age, the greater the chance of death (median age of deaths 79.77 years and for survivors 51.38 years) (Figure 1).

In the assessment of risk factors and outcome between deaths versus survivors, it was observed that chronic cardiovascular disease and diabetes were more frequent in both situations and that those vaccinated against influenza (<0.04) had a higher COVID-19 survival rate (<0.01). ICU admission (<0.02) and invasive ventilation were associated with death (<0.01) (Table 2).

**Table 2.** Risk factors and outcome between deaths versus survivors in SARS by COVID-19 in indigenous people, 03/01/2020 to 02/22/2021, Pará, Brazil.

| Variables | Survivors (77) | % | Death (46) | % | *p-value survivor versus death <sup>(1)</sup> |
| --- | --- | --- | --- | --- | --- |
| Chronic Cardiovascular Disease | 10 | 12.99 | 8 | 17.39 | 0.69 |
| Asthma | 1 | 1.30 | 0 | 0.00 | 0.80 |
| Diabetes mellitus | 7 | 9.09 | 3 | 6.52 | 0.87 |
| Chronic Neurological Disease | 2 | 2.60 | 0 | 0.00 | 0.71 |
| Other Chronic Lung Disease | 1 | 1.30 | 0 | 0.00 | 0.80 |
| Immunodeficiency / Immunodepression | 0 | 0.00 | 2 | 4.35 | 0.28 |
| Chronic Kidney Disease | 2 | 2.60 | 2 | 4.35 | 1.00 |
| Obesity | 2 | 2.60 | 1 | 2.17 | 0.65 |
| Vaccinated against influenza | 24 | 31.17 | 6 | 13.04 | $<0.04$ |
| admitted to the ICU | 9 | 11.69 | 14 | 30.43 | $<0.02$ |
| Invasive ventilation | 3 | 3.90 | 11 | 23.91 | $<0.01$ |

<sup>1</sup> Test G. Source: OpenDataSUS, Ministry of Health, Brazil.

In multivariate logistic regression with the dependent variable death associated with the other variables, adjusted (*log likelihood 124.9322-<0.01). It was shown that invasive ventilation presents (10,73) more chances for the outcome death. On the other hand, those not vaccinated against influenza presented (3.41) more chances of death. Followed by age (1.4) chances of death. ICU and gender were not significant in the model and were removed. (Table 3).

**Table 3.** Odds ratio for death in SARS by COVID-19 in indigenous people, 03/01/2020 to 02/22/2021, Pará, Brazil.

| Variables | Coefficient | Standard error | Z | p-value | odds ratio | Confidence Interval 95% |
| --- | --- | --- | --- | --- | --- | --- |
| Invasive Ventilation | 2,37 | 0.79 | 2.99 | 0.00 | 10.73 | 2.27 a 50.80 |
| Not vaccinated against the flu | 1.23 | 0.56 | 2.18 | 0.02 | 3.41 | 1.13 a 10.24 |
| Age | 0.04 | 0.01 | 4.02 | <0.01 | 1.04 | 1.02 a 1.05 |

Multivariate: $2 * \log(\text{likelihood})$ For this model: 126.6017 Only intercept: 162.6163. Chi-square: 36.0146. G freedom: 3. Probability: < 0.0001. Equation: $\text{Logit Pi} = -3.6825 + (1.2259 X1) + (2.3733 X2) + (0.00353 X3)$ . Source: OpenDataSUS, Ministry of Health, Brazil.

There were reports of cases of SARS by COVID-19 in indigenous people, in 30 of the 144 (20.83%) municipalities in the state, and the municipalities with the highest occurrence of cases were Altamira (17-13.82%) and Novo Repartimento (15-12.19%). These municipalities make up the Special Indigenous Sanitary Districts (DSEIs) Altamira and Guamá Tocantins respectively (Figure 2).

**Figure 2.**
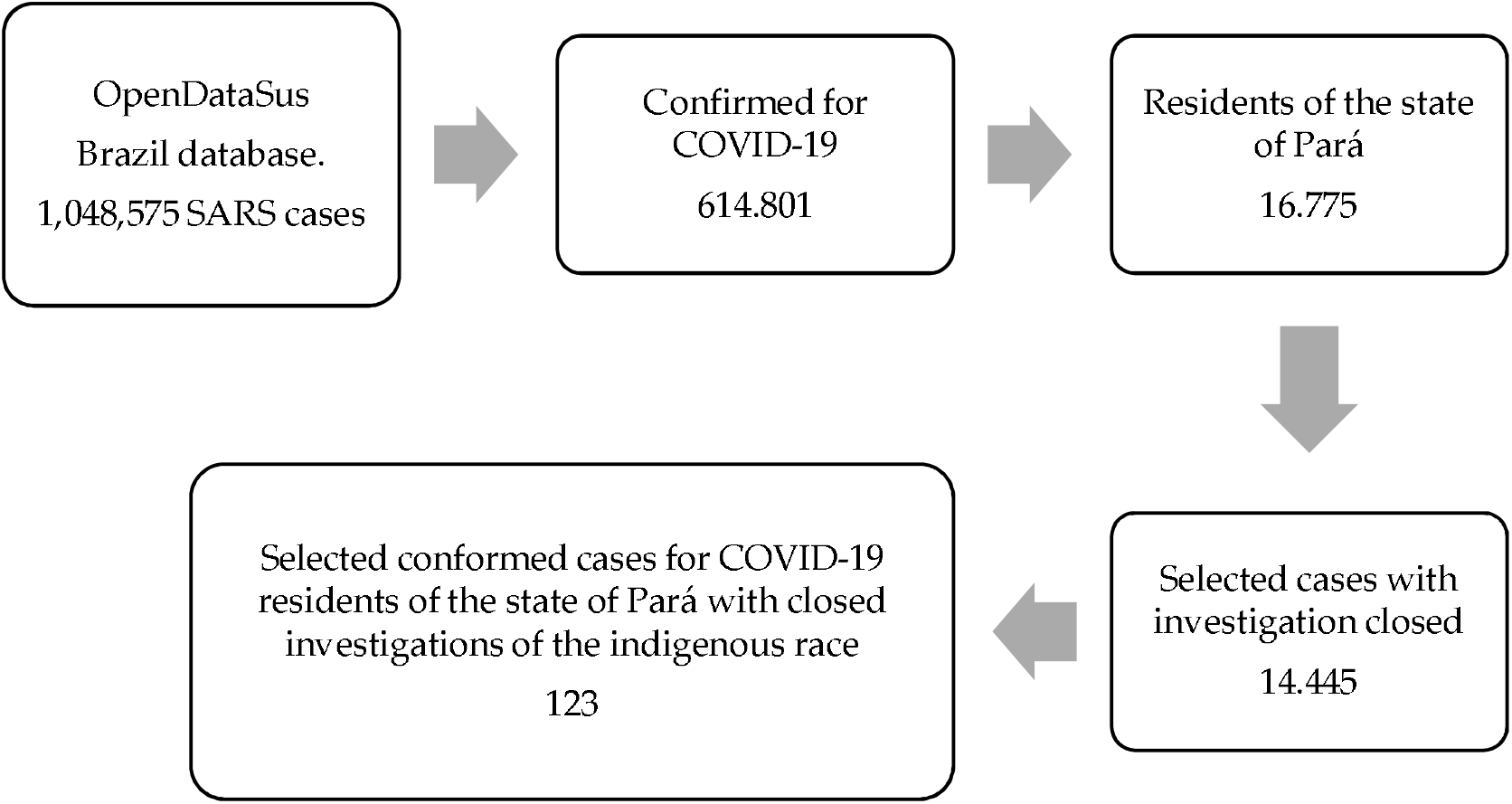
Flowchart of participant selection..

The occurrence of deaths was identified in fifteen municipalities (10.41%), with the highest proportion in the capital Belém (7-15.25), Parauapebas, Santarém and Tucurui (5-10.90%) each (Table 4).

**Table 4.**
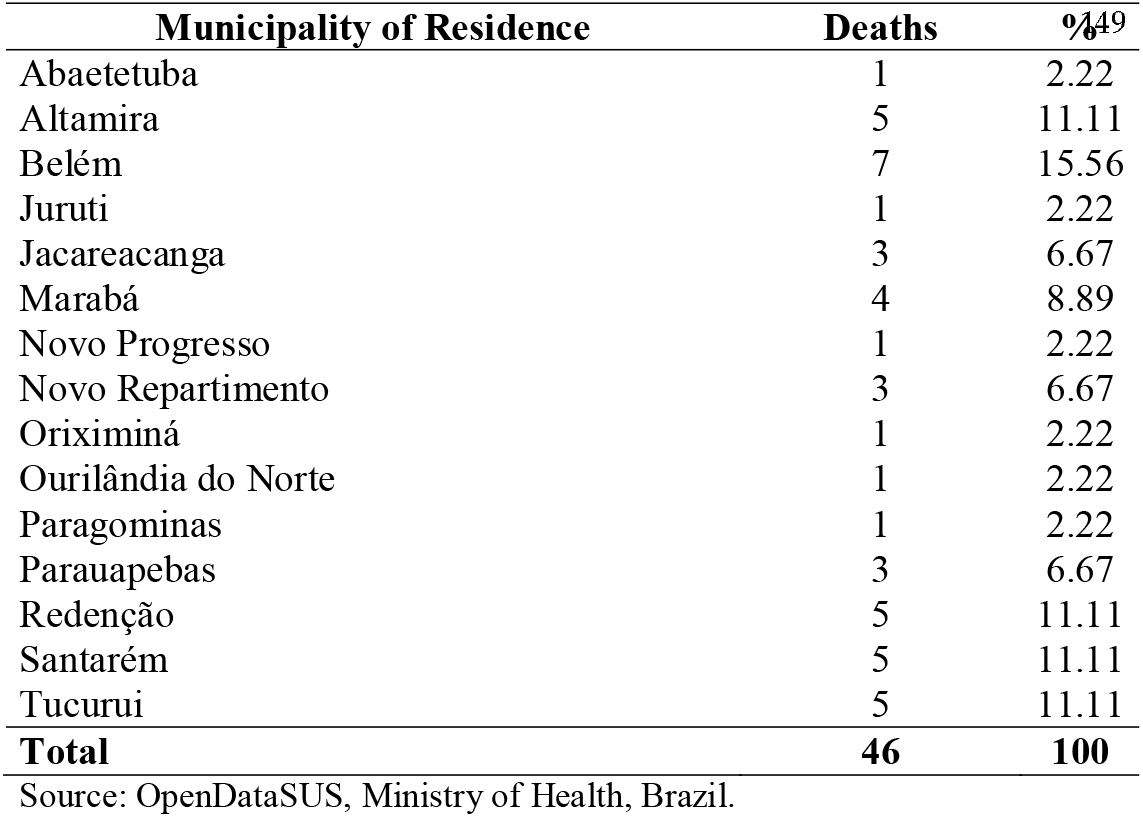
Distribution of deaths from SARS by COVID-19 in indigenous people by municipality of residence, 03/01/2020 to 02/22/2021, Pará, Brazil, 2021.

| Municipality of Residence | Deaths | %<sup>49</sup> |
| --- | --- | --- |
| Abaetetuba | 1 | 2.22 |
| Altamira | 5 | 11.11 |
| Belém | 7 | 15.56 |
| Juruti | 1 | 2.22 |
| Jacareacanga | 3 | 6.67 |
| Marabá | 4 | 8.89 |
| Novo Progresso | 1 | 2.22 |
| Novo Repartimento | 3 | 6.67 |
| Oriximiná | 1 | 2.22 |
| Ourilândia do Norte | 1 | 2.22 |
| Paragominas | 1 | 2.22 |
| Parauapebas | 3 | 6.67 |
| Redenção | 5 | 11.11 |
| Santarém | 5 | 11.11 |
| Tucuruí | 5 | 11.11 |
| **Total** | **46** | **100** |

Source: OpenDataSUS, Ministry of Health, Brazil.

## 4. Discussion

This study analyzed the main epidemiological characteristics of 123 cases and 46 (37.3%) deaths of SARS by COVID-19 in the indigenous population reported by SIVEP-Gripe in the state of Pará from March 1, 2020, to February 22, 2021. It is important to consider that SIVEP-Gripe records and stores only compulsory notifications of hospitalized cases and deaths of SARS by COVID-19 or another etiological agent.

The data showed that males and those over 60 years of age were the most affected. The scientific literature reports that males have presented more severe forms of the disease and consequently evolution to death (Gebhard, Regitz-Zagrosek, Neuhauser, Morgan, & Klein, 2020). Studies try to define the main factors involved in this difference. Among them, that women have a better immune response than men. Pathophysiogenesis studies the receptors of the angiotensin-converting enzyme (ACE2), which is influenced by sex hormones, being the main entry point used by viruses to invade host cells. It is also important to assess differences in gender and social and cultural behaviors, men tend to have higher occurrences of comorbidities, risk behaviors, smoking, etc. The hypothesis of ACE2 receptors is also used to explain higher age-related mortality, as identified in this study (Klein et al., 2020). The elderly would have different expression of ACE2. It is obvious that the elderly are the biggest carriers of chronic diseases (Silva, Rosa, Mendonça, Queiroz, & Oliveira, 2021).

Several researchers corroborate this result, such as European studies where deaths were concentrated up to the epidemiological week 32/2020, in individuals over 65 years old (89%). Deaths in China were more frequent in people over 60 years of age, and also in Mexico. In Brazil, deaths in people over 60 years old accounted for 69.3% of deaths, and of these 64% had at least one risk factor (Barbosa et al., 2020; Escobar, Rodriguez, & Monteiro, 2020)

Studies on the COVID-19 pandemic and indigenous peoples in Brazil showed the same result identified in the population as a whole, however, in the indigenous population this fact represents a threat to the maintenance of these peoples’ cultures, since the elderly are the ones who are holders of these traditional knowledge (Pontes et al., 2021).

It was shown that risk factors for chronic cardiovascular disease (survivors - 12.99% and deaths - 17.39%) and diabetes mellitus (survivors - 9.09% and deaths - 6.52%) were more frequent among cases that evolved to death and to the survivors. A study carried out in Brazil, comparing comorbidities between individuals with SARS due to COVID-19 with the general population, shows that individuals with these comorbidities have a greater chance of evolving to death, therefore, a greater need for hospitalization for the disease (Niquini et al., 2020).

Being corroborated by the study (Sardinha, Lima, et al., 2021) which identified chronic heart diseases (18.97%) and diabetes (18.97%) among the most prevalent comorbidities in the indigenous population with SARS due to COVID-19.

It is important to reflect that the most frequent morbidities among indigenous people are communicable diseases, but due to the urbanization process and change in lifestyle, chronic non-communicable diseases (NCDs) are being identified with increasing frequency in this population (Stein, 2018).

Regarding the diagnostic confirmation criteria, 90.24% of the indigenous people in this study had diagnosis confirmation by identifying the SARS-CoV-2 virus by reverse transcription polymerase chain reaction (RT-PCR), showing that, even in the face of the difficulties, the health services used the gold standard diagnosis, a very sensitive and specific technique, indicated by the World Health Organization (WHO) (Nataraj & Ingole, 2020).

In the analysis for the variables most likely to result in death, the results identified significance for not vaccinated against influenza (p-value < 0.04), ICU admission (p-value < 0.02) and use of invasive ventilation (p-value < 0.01). These results corroborate the literature. (Sardinha et al., 2020) also report greater survival by SRAG among those vaccinated against influenza in Brazil.

Retrospective cohort studies conducted by the University of Michigan (Conlon, Ashur, Washer, Eagle, & Bowman, 2021) identified a significant reduction (24%) in the possibility of testing positive for COVID-19 in individuals who received a dose of influenza vaccine compared with those who did not receive the vaccine. They also concluded that patients previously vaccinated against influenza, with a positive test for COVID-19, had a better clinical outcome, being less likely to be hospitalized or mechanically ventilated and, consequently, had a shorter hospital stay. Similarly, in a study conducted in Brazil with 472,688 severe cases, the independence test was highly significant in vaccinated survivors (<0.0001), and the binary regression showed an odds ratio almost twice as high for invasive ventilation, admission to the hospital. ICU and death in unvaccinated cases (Sardinha, Lobato, et al., 2021).

Brazilian indigenous peoples are among the priority groups for immunization against the influenza virus from six months of age. According to the vacinometer provided by the MS, 77.20% of the indigenous people in the state of Pará received the influenza vaccine in 2020 (M. da S. Brasil, 2021).

A study regarding the characteristics of SARS by COVID-19 in indigenous people in Brazil also identified that being hospitalized in the ICU is a predictor of risk of death, with a chance of 3.96 (OR 3.960: CI 2.913-5,383: <0.0001) (Sardinha, Lima, et al., 2021).

There was a greater proportion of cases and deaths in municipalities that have an indigenous population, or reference services for this population. In addition, there is a greater flow of indigenous people in places where there is a concentration of services and commerce, intensifying this group’s vulnerability. Although indigenous people receive basic care in the villages, more complex services only take place in urban areas (Silva et al., 2021).

This study did not separately assess indigenous people living in villages from those living outside the villages, in this respect, municipalities in the state such as Belém and Santarém received a large number of indigenous people of the Warao ethnic group from Venezuela who live in shelters administered by the state government and by city halls in situations of intense vulnerability to the spread of the virus.

## 5. Conclusions

It was possible to evidence the epidemiological characteristics of severe cases and deaths from 123 COVID-19 cases in the indigenous population based on data from SIVEP-GRIPE, which are from mainly male, over 60 years old. The most prevalent comorbidities are chronic cardiovascular disease and diabetes mellitus. It has also been shown that non-influenza vaccination, admission to the ICU, and use of invasive ventilation increase the chances of death.

Severe cases and deaths are distributed in 30 and 15 municipalities, respectively. However, the occurrence was higher in municipalities that have villages or are references for health care or commerce, in addition to those that received migrants from the Warao ethnic group.

Since the indigenous people have factors of greater vulnerability to the disease and difficulties in accessing more complex health services in the event of the severity of the condition, it is extremely important that the data on death by COVID-19 be updated, considering the occurrence of SRAG and deaths that remained in the villages and were not included in the SIVEP-GRIPE. Notifications must consider the specific issues of indigenous peoples so that effective control measures can be defined.

## Data Availability

https://opendatasus.saude.gov.br/

## Author Contributions

.A.L of the S.F. developed the project, the writing of the article (results and discussion). D.M.S. performed the statistical analyses and contributed to the discussion. C.O.E contributed to writing the introduction, methodology and discussion. C.A.F. de S. performed the data analysis regarding the spatial distribution (map). E.C.C. performed the translation and supervision of the research. K.V.B.L. supervised the research. All authors approved the final version.

## Funding

Conselho Nacional de Desenvolvimento Científico e Tecnológico (CNPQ). Coordenação de Aperfeiçoamento de Pessoal de Nível Superior (CAPES). Fundação Amazônia de Amparo a Estudos e Pesquisas (FAPESPA) and Instituto Evandro Chagas (IEC).

## Acknowledgments

We are grateful for the contribution of the State Secretariat of Health of Pará for data available.

## Conflicts of Interest

The authors declare no conflict of interest.

## Notes

### Competing Interest Statement

The authors have declared no competing interest.

### Funding Statement

CAPES
CNPQ

